# Atovaquone-Proguanil and Reduced Digestive Cancer Risk: A *Toxoplasma gondii* Connection

**DOI:** 10.1101/2025.03.24.25324497

**Authors:** Ariel Israel, Sarah Israel, Abraham Weizman, Shai Ashkenazi, Shlomo Vinker, Eli Magen, Eugene Merzon

**Affiliations:** Leumit Research Institute, Leumit Health Services, 23 Sprinzak St., Tel Aviv, Israel; Department of Epidemiology and Preventive Medicine, School of Public Health, Faculty of Medical & Health Sciences, Tel Aviv University; Ramat Aviv, Tel Aviv, Israel; Department of Clinical Microbiology and Infectious Diseases, Hadassah-Hebrew University Medical Center, Faculty of Medicine, Hebrew University of Jerusalem, P.O. Box 12000, Jerusalem, Israel; Research Unit, Geha Mental Health Center, Felsenstein Medical Research Center, and Sagol School of Neurosciences, Faculty of Medical & Health Sciences, Tel Aviv University, Tel Aviv, Israel; Adelson School of Medicine, Ariel University, Ariel, Israel; Unit of Infectious Diseases, Schneider Children’s Medical Center, 14 Kaplan St., Petah Tikva, Israel; Medicine A Department, Assuta Ashdod University Medical Center, Ben Gurion University of the Negev, Derech HaRofe 7, Ashdod, Israel

**Keywords:** Colorectal Cancer, Pancreatic Cancer, Toxoplasma gondii, Atovaquone, Metagenomics, Microbiota, Cancer Prevention, Electronic Health Records

## Abstract

Emerging evidence suggests microbial pathogens contribute to digestive cancer risk. Atovaquone–proguanil (A-P), an antimalarial with antiparasitic activity, has been associated with a reduced risk of colorectal cancer (CRC). We conducted a retrospective cohort study using the TriNetX US Collaborative Network, including over 100,000 individuals aged 40–69 years who received A-P, matched 1:1 to controls who received other medications. Incident digestive cancers were analyzed using Cox proportional hazards models. Additionally, we performed a metagenomic analysis of 1,044 fecal samples from 156 individuals to assess the abundance of *Toxoplasma gondii* in CRC-associated microbiota. A-P use was associated with a significant reduction in digestive cancer incidence across all age groups: hazard ratios (HRs) ranged from 0.49 to 0.53 (all P<0.001). Protective associations extended to pancreatic cancer (HR range, 0.50–0.72). In metagenomic analysis, *T. gondii* was the most discriminatory microbial species for CRC (P=1.8×10^−16^), detected above threshold in 22.6% of CRC samples versus 1.6% of controls (odds ratio 18.2, 95% CI, 8.2–47.6, P=2.3×10^−22^). These findings suggest *T. gondii* may be an overlooked microbial risk factor for digestive cancers, and that A-P may offer chemopreventive effects through antiparasitic activity. Prospective studies are needed to evaluate its preventive potential.

## Introduction

Digestive tract cancers, including those of the colon, pancreas, and stomach, are among the leading causes of cancer-related deaths worldwide. Colorectal cancer (CRC) alone accounts for over 900,000 deaths annually and remains the second most common cause of cancer mortality globally ^1^. While genetic predisposition, inflammation, and environmental exposures contribute to cancer risk, microbial factors have emerged as important but incompletely understood influences on digestive tract carcinogenesis.^2^

Bacterial pathogens such as *Helicobacter pylori* and *Fusobacterium nucleatum* have been implicated in gastric and colorectal cancers, respectively ^3,4^. However, most studies of the cancer-associated microbiome have focused on bacteria, often overlooking protozoa, which are typically undetectable by 16S rRNA gene sequencing and often absent from metagenomic reference databases^5,6^.

Atovaquone-proguanil (A-P) is a well-tolerated antimalarial drug combination used primarily for short-term prophylaxis. In prior observational studies, A-P exposure has been associated with reduced incidence of several cancers ^7^. Although this has been attributed to its mitochondrial inhibitory effects, such a mechanism does not fully explain the prolonged reduction in cancer risk observed years after treatment. Notably, atovaquone exhibits antiparasitic activity against apicomplexan protozoa such as *Toxoplasma gondii* ^8^ through inhibition of their mitochondrial electron transport chain^9^, suggesting that eradication of such pathogens may underlie its long-term anticancer effect.

*T. gondii*, a widespread protozoan parasite,^10^ has been linked primarily to neurological and ocular diseases in immunocompromised or congenitally infected individuals^11^. However, *T. gondii* also exhibits a strong tropism for the intestinal epithelium during both acute and chronic infection, where it can persist within tissue cysts and elicit chronic, low-grade inflammation^12^. This mucosal colonization induces local immune activation—characterized by elevated IL-6 and TNF-α—and disrupts epithelial barrier integrity, potentially creating a pro-oncogenic environment^13^. Despite this, its role in gastrointestinal malignancies has not been systematically explored.

To investigate this possibility, we employed a dual analytic approach. First, we conducted a retrospective cohort analysis using a large electronic health record network to assess associations between A–P use and long-term digestive cancer risk. Second, we reanalyzed public metagenomic sequencing data using a protozoa-inclusive reference database to evaluate the prevalence of *T. gondii* in the fecal microbiota of patients with and without colorectal cancer. To our knowledge, this is one of the first studies to integrate large-scale electronic health record data and metagenomic profiling to assess protozoan involvement in gastrointestinal cancer risk.

## Materials and Methods

### Study Design and Data Sources

We conducted a two-part investigation to evaluate the association between atovaquone-proguanil (A-P) use, *Toxoplasma gondii* presence, and digestive cancer risk. The first part was a retrospective cohort analysis using the TriNetX US Collaborative Network, a federated health research platform aggregating deidentified electronic health records from 70 healthcare organizations^14^, including over 120 million patients as of March 2025. The second part involved a reanalysis of publicly available metagenomic shotgun sequencing data (PRJEB6070) to assess *T. gondii* prevalence in colorectal cancer (CRC)-associated microbiota. PRJEB6070 consists of fecal samples from 156 individuals in France who underwent colonoscopy to diagnose colorectal neoplasia (adenomas or CRC) or confirm its absence^15^.

### TriNetX Cohort Analysis

We created three propensity score-matched (PSM) cohorts of individuals aged 40-49, 50-59, and 60-69 years who received A-P compared to those who received any other medication. Exposure in the A–P group was defined as a recorded medication purchase that included both atovaquone and proguanil, occurring within the age-defined interval. The comparator groups consisted of individuals with any recorded medication purchase within the same age-defined interval, with the sole exclusion criterion being any recorded use of atovaquone, to minimize contamination of the control group. These control groups were designed to represent individuals utilizing pharmacy services under circumstances similar to those of the A–P group, but without exposure to the antiparasitic agent. Control participants were matched 1:1 using logistic regression on age, sex, race, smoking status, body mass index, and diabetes status. Individuals with prior diagnoses of the outcome of interest were excluded from the analyses. Follow-up extended from the index date (first recorded medication) to outcome diagnosis, death, or last available clinical record. Hazard ratios (HRs) were estimated using Cox proportional hazards models, with Kaplan-Meier curves and Schoenfeld residuals used to test proportionality.

### Metagenomic Analysis

We reanalyzed metagenomic sequencing data from 1,044 fecal samples from PRJEB6070. Taxonomic classification was performed using Centrifuge (v1.0.4)^16^ with the SANBAFPH reference database, which includes bacterial, fungal, protozoan, nematode, and human genomes^17^. Centrifuge outputs the number of reads uniquely mapped to each taxon, which we used as a semi-quantitative measure of organismal load across samples. Although Centrifuge does not compute normalized abundance estimates (e.g., accounting for genome size or total sequencing depth), the consistent DNA extraction and sequencing protocols across all samples support valid comparative analysis based on raw read counts.

We selected Centrifuge over other popular tools such as MetaPhlAn, as MetaPhlAn is designed primarily for bacterial profiling using clade-specific marker genes and does not support protozoan classification^18^. In contrast, Centrifuge permits broad detection of microbial eukaryotes when paired with an appropriate reference database.

Taxonomic classification was based on best match, with sequences assigned to an organism if they uniquely aligned with ≥95% identity. For each organism, person-level detection was defined as the average number of uniquely mapped reads across all available samples for that individual (approximately six samples per individual). For *Toxoplasma gondii*, presence was further evaluated using an empirical threshold of ≥50 uniquely mapped reads averaged across the individual’s samples.

### Statistical Analyses

In the TriNetX cohort analysis, we estimated HRs with 95% confidence intervals (CIs) using Cox proportional hazards models. We analyzed digestive malignancies as a composite outcome (ICD-10 C15–C26), and also conducted a subgroup analysis for pancreatic cancer (ICD-10 C25). To improve internal validity and assess consistency across life stages, we stratified the A–P and control cohorts by decade (ages 40–49, 50–59, and 60–69) before matching. Each age-stratified cohort was constructed independently using TriNetX’s “compare outcomes” framework, and the analysis was performed separately for the digestive cancer and pancreatic cancer analyses. As a result, baseline demographic characteristics are closely matched but not identical across cohorts, and sample sizes and outcome counts differ between analyses, as reflected in Table 1.

**Table 1:**
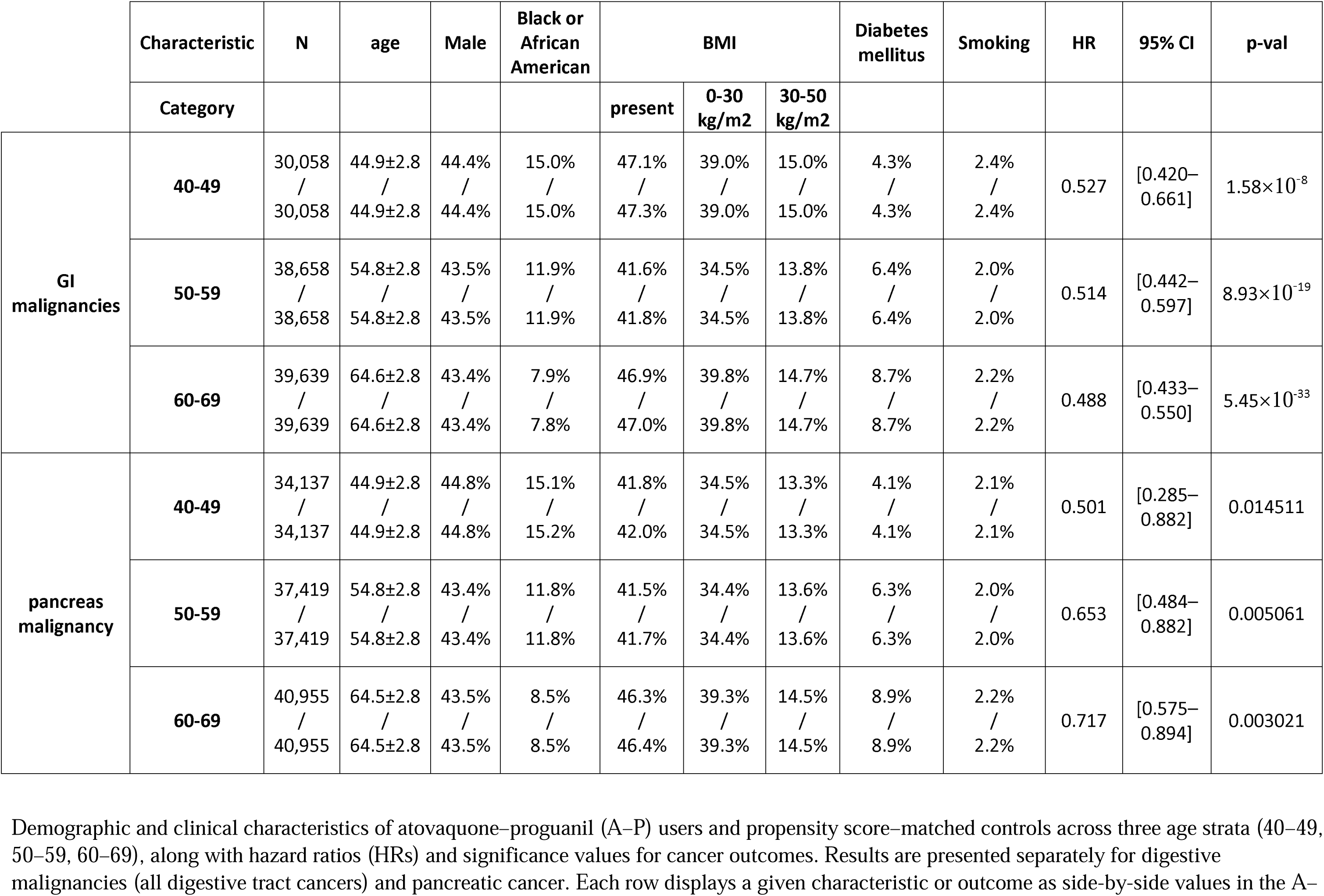
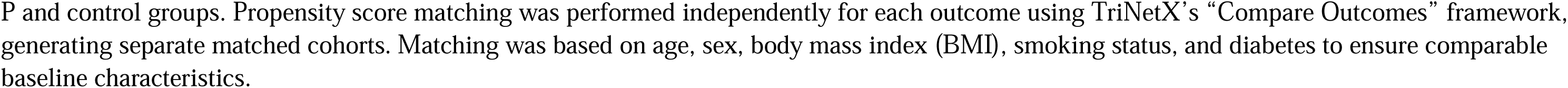
Statistics of the TriNetX Cohorts.

|  | Characteristic | N | age | Male | Black or African American | BMI |  |  | Diabetes mellitus | Smoking | HR | 95% CI | p-val |
| --- | --- | --- | --- | --- | --- | --- | --- | --- | --- | --- | --- | --- | --- |
|  | Category |  |  |  |  | present | 0-30 kg/m2 | 30-50 kg/m2 |  |  |  |  |  |
| GI malignancies | 40-49 | 30,058<br>/<br>30,058 | 44.9±2.8<br>/<br>44.9±2.8 | 44.4%<br>/<br>44.4% | 15.0%<br>/<br>15.0% | 47.1%<br>/<br>47.3% | 39.0%<br>/<br>39.0% | 15.0%<br>/<br>15.0% | 4.3%<br>/<br>4.3% | 2.4%<br>/<br>2.4% | 0.527 | [0.420–0.661] | 1.58×10<sup>-8</sup> |
|  | 50-59 | 38,658<br>/<br>38,658 | 54.8±2.8<br>/<br>54.8±2.8 | 43.5%<br>/<br>43.5% | 11.9%<br>/<br>11.9% | 41.6%<br>/<br>41.8% | 34.5%<br>/<br>34.5% | 13.8%<br>/<br>13.8% | 6.4%<br>/<br>6.4% | 2.0%<br>/<br>2.0% | 0.514 | [0.442–0.597] | 8.93×10<sup>-19</sup> |
|  | 60-69 | 39,639<br>/<br>39,639 | 64.6±2.8<br>/<br>64.6±2.8 | 43.4%<br>/<br>43.4% | 7.9%<br>/<br>7.8% | 46.9%<br>/<br>47.0% | 39.8%<br>/<br>39.8% | 14.7%<br>/<br>14.7% | 8.7%<br>/<br>8.7% | 2.2%<br>/<br>2.2% | 0.488 | [0.433–0.550] | 5.45×10<sup>-33</sup> |
| pancreas malignancy | 40-49 | 34,137<br>/<br>34,137 | 44.9±2.8<br>/<br>44.9±2.8 | 44.8%<br>/<br>44.8% | 15.1%<br>/<br>15.2% | 41.8%<br>/<br>42.0% | 34.5%<br>/<br>34.5% | 13.3%<br>/<br>13.3% | 4.1%<br>/<br>4.1% | 2.1%<br>/<br>2.1% | 0.501 | [0.285–0.882] | 0.014511 |
|  | 50-59 | 37,419<br>/<br>37,419 | 54.8±2.8<br>/<br>54.8±2.8 | 43.4%<br>/<br>43.4% | 11.8%<br>/<br>11.8% | 41.5%<br>/<br>41.7% | 34.4%<br>/<br>34.4% | 13.6%<br>/<br>13.6% | 6.3%<br>/<br>6.3% | 2.0%<br>/<br>2.0% | 0.653 | [0.484–0.882] | 0.005061 |
|  | 60-69 | 40,955<br>/<br>40,955 | 64.5±2.8<br>/<br>64.5±2.8 | 43.5%<br>/<br>43.5% | 8.5%<br>/<br>8.5% | 46.3%<br>/<br>46.4% | 39.3%<br>/<br>39.3% | 14.5%<br>/<br>14.5% | 8.9%<br>/<br>8.9% | 2.2%<br>/<br>2.2% | 0.717 | [0.575–0.894] | 0.003021 |

Demographic and clinical characteristics of atovaquone–proguanil (A–P) users and propensity score–matched controls across three age strata (40–49, 50–59, 60–69), along with hazard ratios (HRs) and significance values for cancer outcomes. Results are presented separately for digestive malignancies (all digestive tract cancers) and pancreatic cancer. Each row displays a given characteristic or outcome as side-by-side values in the A–
P and control groups. Propensity score matching was performed independently for each outcome using TriNetX's "Compare Outcomes" framework, generating separate matched cohorts. Matching was based on age, sex, body mass index (BMI), smoking status, and diabetes to ensure comparable baseline characteristics.

This approach enabled comparison within more homogeneous age groups, ensured that the observed associations were not restricted to a single age category, and allowed us to test the persistence of the protective association at three different points in the adult life course. Given the limited long-term follow-up in TriNetX, such stratification strengthens causal plausibility by demonstrating consistent associations at distinct index ages.

In the metagenomic analysis, we first calculated the mean number of reads per organism, per individual, across all available fecal samples. Statistical comparisons between individuals with and without colorectal cancer (CRC) were then performed using the Mann–Whitney U test on these per-individual read count averages. Microbial species were ranked by p-value, with *T. gondii* emerging as the most discriminatory taxon out of 8,425 detected. We then applied an empirical threshold of ≥50 uniquely mapped reads to define *T. gondii* presence and used Fisher’s exact test to evaluate its association with CRC status. Diagnostic performance was further assessed using receiver operating characteristic (ROC) curve analysis. Mixed-effects models were not employed, as our analysis was based on per-individual summary values rather than repeated measures per sample.

All statistical analyses were conducted using Python (version 3.11.9) and R (version 4.4.0), with two-tailed P values <0.05 considered statistically significant.

### Ethics Statement

The study was approved by the Leumit Health Services Institutional Review Board (approval number LEU-0010-21). Informed consent was waived due to the use of deidentified data. The metagenomic dataset (PRJEB6070) is publicly available through the European Nucleotide Archive.

## Results

### TriNetX Cohort Analyses

In the propensity score-matched TriNetX cohorts, atovaquone-proguanil (A-P) use was associated with a significantly lower incidence of digestive cancers (ICD-10 C15-C26) across all age groups (Table 1, Figure 1).

- In the 40-49 age group (mean age, 44.9 years; n = 30,058 per group), A-P use was associated with a **47% lower risk** of digestive cancer compared to controls (hazard ratio [HR], 0.53; 95% CI, 0.42-0.66).
- In the 50-59 age group (mean age, 54.8 years; n = 38,658 per group), the risk reduction was **49%** (HR, 0.51; 95% CI, 0.44-0.60).
- In the 60-69 age group (mean age, 64.6 years; n = 39,643 per group), the reduction was **51%** (HR, 0.49; 95% CI, 0.43-0.55).

**Figure 1:**
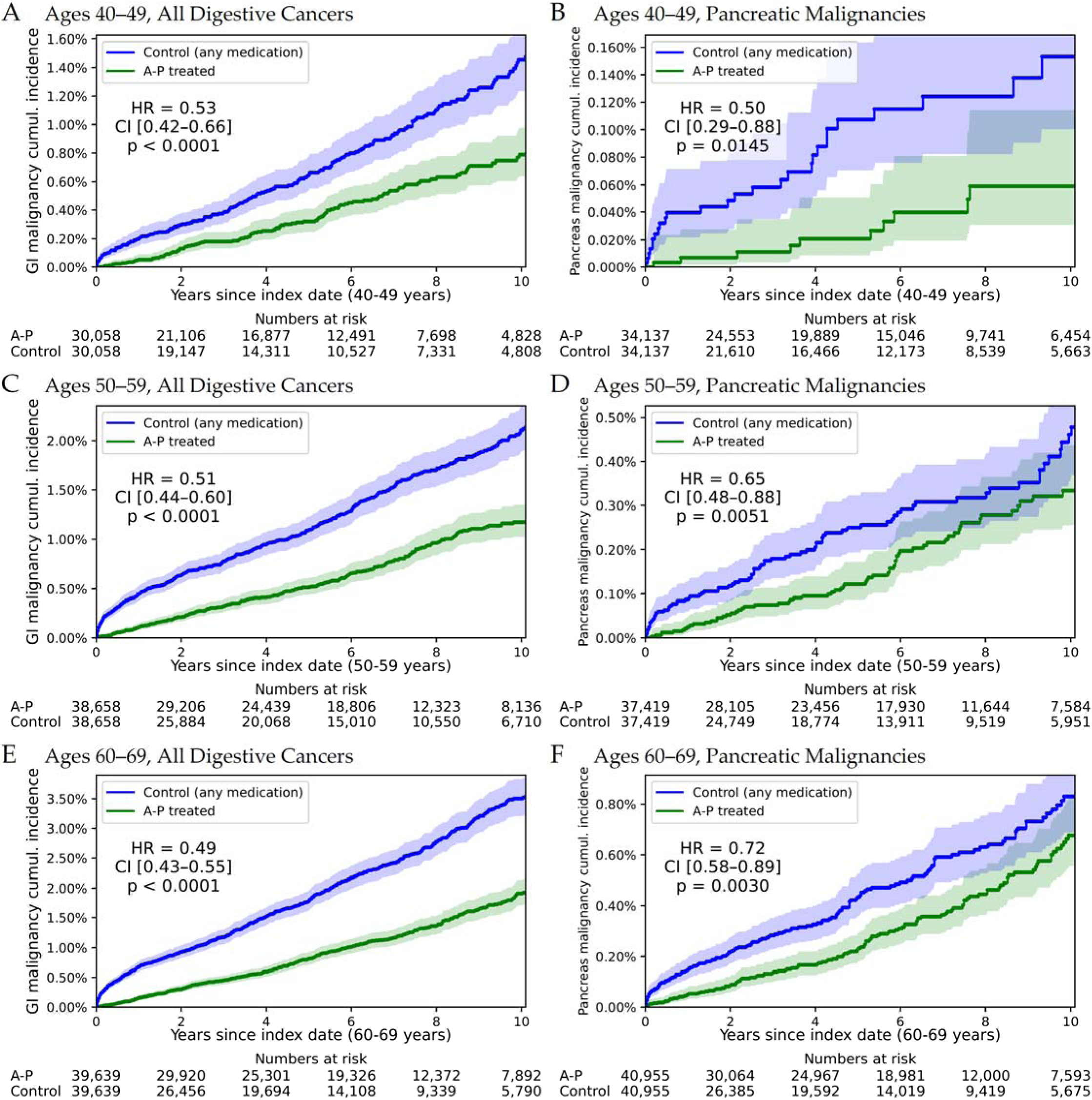
Cumulative Incidence of Digestive Cancers and Pancreatic Malignancies in Propensity Score-Matched TriNetX Cohorts. Kaplan–Meier curves showing the cumulative incidence of digestive cancers (ICD-10 codes C15–C26) and pancreatic malignancies among users of atovaquone–proguanil (A–P, blue) versus propensity score–matched controls (green) across three age cohorts. Shaded areas represent 95% confidence intervals. Numbers at risk are presented at 2-year intervals. P-values were derived from log-rank tests.

Colorectal cancer (ICD-10 C18–C20), the most frequent digestive tract malignancy in this cohort, showed a similarly strong association. In the 60–69 age group, the hazard ratio was 0.44 (95% CI, 0.36–0.54), reflecting a 55% reduction in risk. This suggests that much of the observed decrease in overall digestive cancer incidence was driven by reduced CRC rates.

### Pancreatic Cancer Analyses

Pancreatic cancer (ICD-10 C25) was analyzed as a specific outcome. A-P use was associated with reduced pancreatic cancer risk across all age groups:

- **40-49 years** (HR, 0.50; 95% CI, 0.29-0.88; n = 34,137 per group),
- **50-59 years** (HR, 0.65; 95% CI, 0.48-0.88; n = 37,419 per group),
- **60-69 years** (HR, 0.72; 95% CI, 0.58-0.89; n = 40,955 per group).

Kaplan-Meier analyses supported a consistent protective association, with less pronounced separation compared to other digestive cancers (Figure 1B, 1D, 1F).

### Metagenomic Analysis

We reanalyzed 1,044 fecal microbiota samples from 156 individuals in the PRJEB6070 dataset, including 53 colorectal cancer (CRC) patients, 42 adenoma patients (27 with small adenomas and 15 with large adenomas), and 61 individuals with normal findings. Cancer patients were slightly older (mean age, 66.8 years) than those without cancer (mean age, 60.6 years), while body mass index did not differ significantly across groups (P = 0.53).

Using Centrifuge with the SANBAFPH database, we detected 8,425 microbial species, with *T. gondii* emerging as the most discriminatory taxon for CRC (Mann-Whitney U test: P = 1.8×10 ¹), ranking above the leading bacterial association, *Fusobacterium* (P = 5.6×10 ¹).

- *T. gondii* (≥50 uniquely mapped reads) was detected in 22.1% of CRC samples compared to 1.53% of normal samples (Fisher’s exact test; odds ratio [OR], 18.20; 95% CI, 8.2-47.6; P = 2.3×10 ²²).
- At the individual level, *T. gondii* presence was observed in 22.5% of CRC patients versus 1.64% of non-cancer individuals (OR, 17.2; 95% CI, 2.4-760.0; P < 0.001).

Cancer patients had significantly higher *T. gondii* read counts than controls (Table 2; Figure 2A), with a dose-response relationship (Figure 2B).

- 15.1% of CRC patients had 50-999 reads compared to 1.6% of controls, and
- 7.6% of CRC patients had >1,000 reads compared to none in controls.
- The relative abundance of *T. gondii* was also significantly elevated in CRC samples.

**Figure 2:**
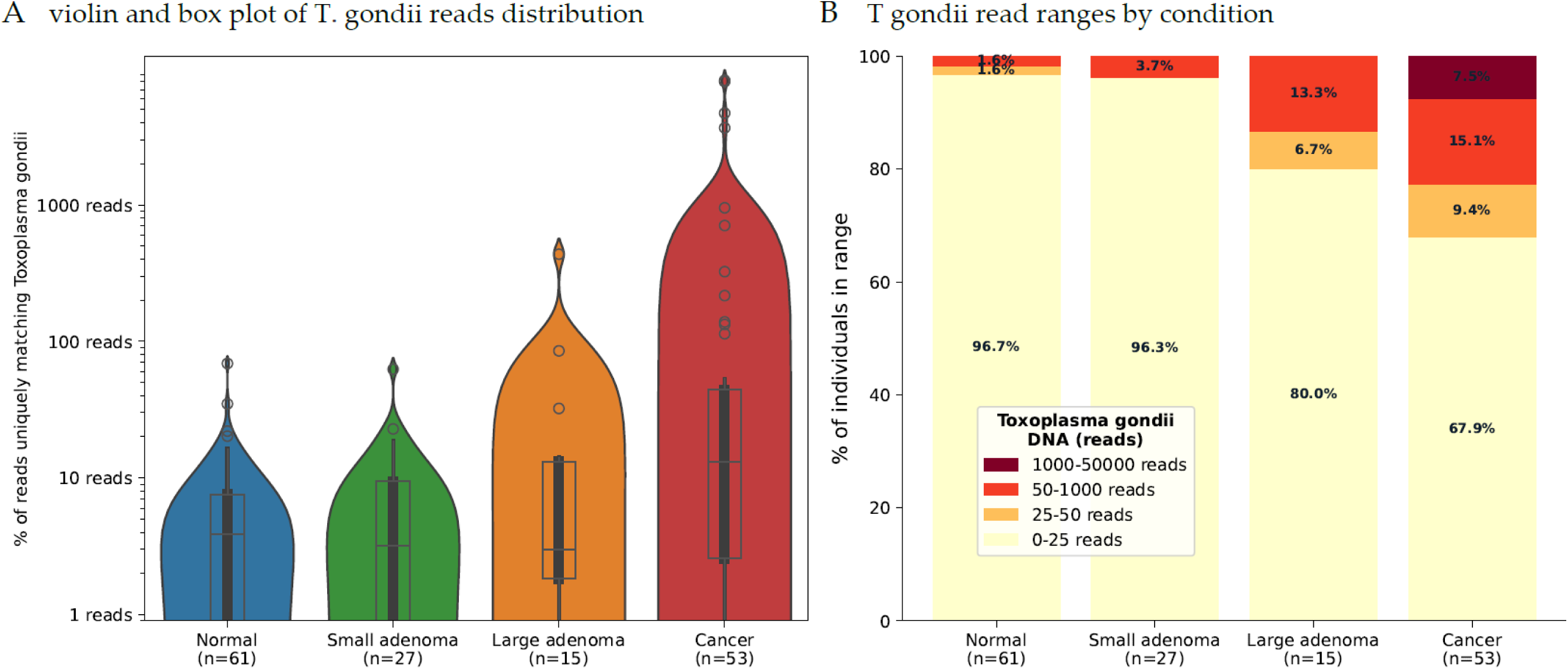
Distribution of *Toxoplasma gondii* Reads by Patient Condition. **Panel A:** Violin and box plots showing the distribution of *T. gondii* reads (log scale) in 1,044 fecal samples from 156 individuals in the PRJEB6070 dataset, grouped by diagnosis: Normal (n = 61), Small Adenoma (n = 27), Large Adenoma (n = 15), and Colorectal Cancer (n = 53). CRC patients exhibited significantly higher *T. gondii* levels, with a progressive increase from normal to cancer (P < 0.001). **Panel B:** Stacked bar plot illustrating the proportion of individuals with mean *T. gondii* read counts in four ranges (0-25, 25-50, 50-1,000, and >1,000 reads) across diagnostic categories.

**Table 2:**
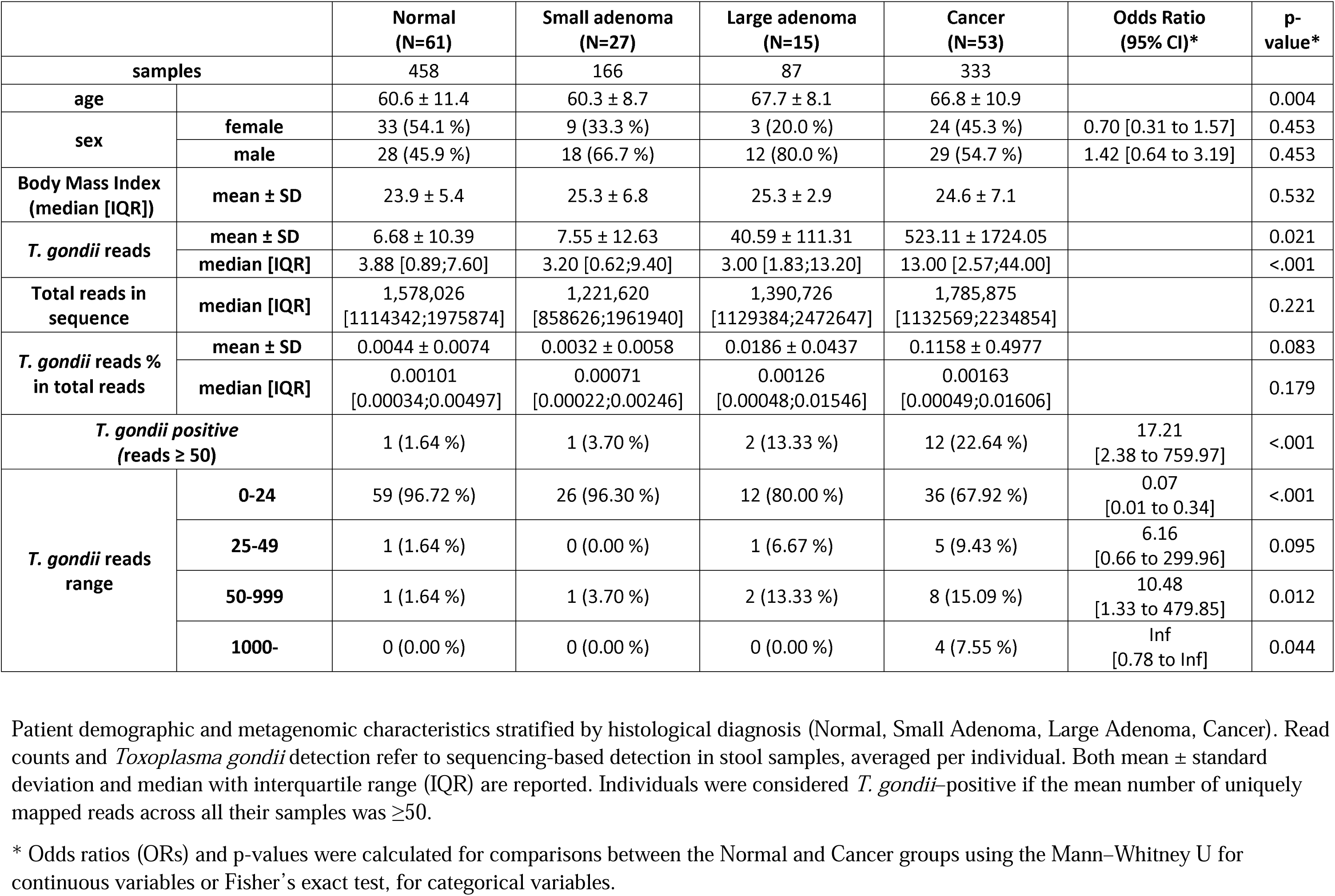
Patient Characteristics and Metagenomic Data by Diagnosis.

|  |  | Normal<br>(N=61) | Small adenoma<br>(N=27) | Large adenoma<br>(N=15) | Cancer<br>(N=53) | Odds Ratio<br>(95% CI)* | p-value* |
| --- | --- | --- | --- | --- | --- | --- | --- |
| **samples** |  | 458 | 166 | 87 | 333 |  |  |
| **age** |  | 60.6 ± 11.4 | 60.3 ± 8.7 | 67.7 ± 8.1 | 66.8 ± 10.9 |  | 0.004 |
| **sex** | **female** | 33 (54.1 %) | 9 (33.3 %) | 3 (20.0 %) | 24 (45.3 %) | 0.70 [0.31 to 1.57] | 0.453 |
|  | **male** | 28 (45.9 %) | 18 (66.7 %) | 12 (80.0 %) | 29 (54.7 %) | 1.42 [0.64 to 3.19] | 0.453 |
| **Body Mass Index**<br>**(median [IQR])** | **mean ± SD** | 23.9 ± 5.4 | 25.3 ± 6.8 | 25.3 ± 2.9 | 24.6 ± 7.1 |  | 0.532 |
| ***T. gondii* reads** | **mean ± SD** | 6.68 ± 10.39 | 7.55 ± 12.63 | 40.59 ± 111.31 | 523.11 ± 1724.05 |  | 0.021 |
|  | **median [IQR]** | 3.88 [0.89;7.60] | 3.20 [0.62;9.40] | 3.00 [1.83;13.20] | 13.00 [2.57;44.00] |  | <.001 |
| **Total reads in**<br>**sequence** | **median [IQR]** | 1,578,026<br>[1114342;1975874] | 1,221,620<br>[858626;1961940] | 1,390,726<br>[1129384;2472647] | 1,785,875<br>[1132569;2234854] |  | 0.221 |
| ***T. gondii* reads %**<br>**in total reads** | **mean ± SD** | 0.0044 ± 0.0074 | 0.0032 ± 0.0058 | 0.0186 ± 0.0437 | 0.1158 ± 0.4977 |  | 0.083 |
|  | **median [IQR]** | 0.00101<br>[0.00034;0.00497] | 0.00071<br>[0.00022;0.00246] | 0.00126<br>[0.00048;0.01546] | 0.00163<br>[0.00049;0.01606] |  | 0.179 |
| ***T. gondii* positive**<br>**(reads ≥ 50)** |  | 1 (1.64 %) | 1 (3.70 %) | 2 (13.33 %) | 12 (22.64 %) | 17.21<br>[2.38 to 759.97] | <.001 |
| ***T. gondii* reads**<br>**range** | **0-24** | 59 (96.72 %) | 26 (96.30 %) | 12 (80.00 %) | 36 (67.92 %) | 0.07<br>[0.01 to 0.34] | <.001 |
|  | **25-49** | 1 (1.64 %) | 0 (0.00 %) | 1 (6.67 %) | 5 (9.43 %) | 6.16<br>[0.66 to 299.96] | 0.095 |
|  | **50-999** | 1 (1.64 %) | 1 (3.70 %) | 2 (13.33 %) | 8 (15.09 %) | 10.48<br>[1.33 to 479.85] | 0.012 |
|  | **1000-** | 0 (0.00 %) | 0 (0.00 %) | 0 (0.00 %) | 4 (7.55 %) | Inf<br>[0.78 to Inf] | 0.044 |

Patient demographic and metagenomic characteristics stratified by histological diagnosis (Normal, Small Adenoma, Large Adenoma, Cancer). Read counts and *Toxoplasma gondii* detection refer to sequencing-based detection in stool samples, averaged per individual. Both mean ± standard deviation and median with interquartile range (IQR) are reported. Individuals were considered *T. gondii*-positive if the mean number of uniquely mapped reads across all their samples was ≥50.
\* Odds ratios (ORs) and p-values were calculated for comparisons between the Normal and Cancer groups using the Mann–Whitney U for continuous variables or Fisher’s exact test, for categorical variables.

### Screening Potential

We evaluated the potential of *T. gondii* read counts as a non-invasive biomarker for colorectal cancer (CRC). Receiver operating characteristic (ROC) curve analysis yielded an area under the curve (AUC) of 0.720 (Figure 3), indicating moderate discriminatory power. For context, a recent meta-analysis of 31 studies reported an AUC of 0.77 (95% CI: 0.75–0.79) for traditional guaiac-based fecal occult blood tests (gFOBT), and a higher AUC of 0.87 (95% CI: 0.85–0.88) for immunochemical FOBTs (iFOBT). While the performance of *T. gondii* read counts is below that of iFOBT, it is comparable to gFOBT, supporting its potential utility as a microbiome-based biomarker for CRC detection, particularly when combined with other features or screening modalities.^19^

**Figure 3:**
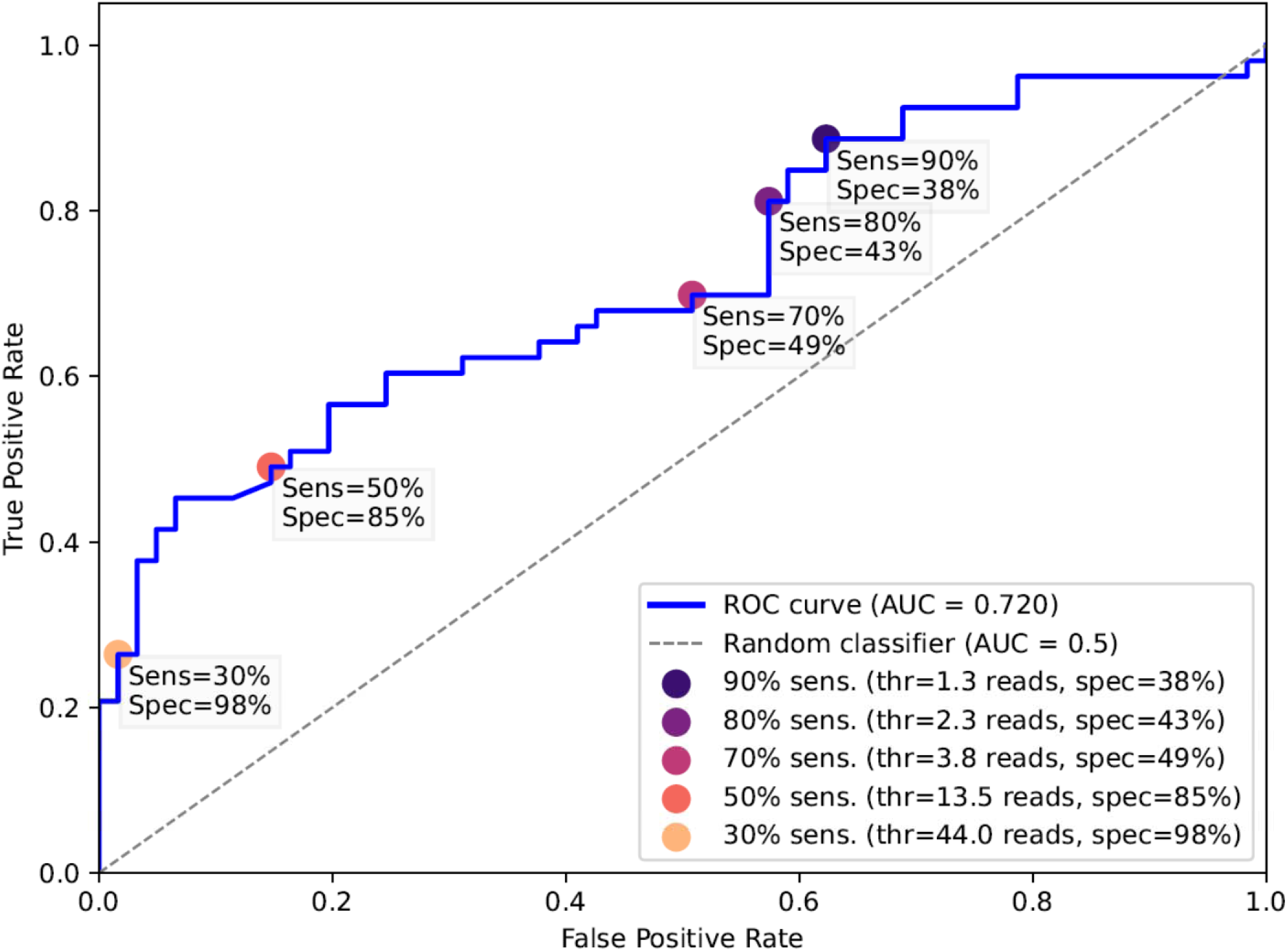
Receiver Operating Characteristic (ROC) Curve for *Toxoplasma gondii* Read Counts in Colorectal Cancer Classification. ROC curve evaluating the ability of T. gondii read counts to distinguish colorectal cancer (CRC) from non-cancer cases in fecal metagenomic samples from 156 individuals in the PRJEB6070 dataset. The area under the curve (AUC) was 0.720, indicating promising discriminatory performance. Selected thresholds are marked on the curve:

- **1.3 reads:** 90% sensitivity, 38% specificity
- **2.3 reads:** 80% sensitivity, 43% specificity
- **3.8 reads:** 70% sensitivity, 49% specificity
- **13.5 reads:** 50% sensitivity, 85% specificity
- **44.0 reads:** 30% sensitivity, 98% specificity These results suggest that *T. gondii* read abundance may serve as a potential biomarker for CRC screening.

Sensitivity and specificity at selected read count thresholds were:

- 90% sensitivity and 38% specificity at 1.3 reads,
- 80% sensitivity and 43% specificity at 2.3 reads,
- 70% sensitivity and 49% specificity at 3.8 reads,
- 50% sensitivity and 85% specificity at 13.5 reads,
- 30% sensitivity and 98% specificity at 44.0 reads.

These findings suggest that *T. gondii* DNA detection using optimized molecular assays may have potential as a screening tool for CRC, particularly at higher thresholds where specificity is high.

## Discussion

In this study, we show—using large, propensity score-matched cohorts from the TriNetX database—that atovaquone–proguanil (A–P) use is associated with a substantial reduction in colorectal cancer (CRC) risk. This association was highly statistically significant and contributed to an overall 47–51% reduction in the incidence of digestive cancers across three age groups (40-49, 50-59, and 60-69 years). In addition, pancreatic cancer risk was reduced by 28–49%, suggesting that A–P may confer protection across a broader spectrum of digestive tract malignancies. The association with pancreatic cancer is particularly noteworthy given its distinct pathogenesis, poor prognosis, and the absence of effective treatment and screening tools.

The sustained divergence of Kaplan–Meier curves more than a decade after treatment supports a durable protective effect, likely through elimination of a long-lived pathogen rather than a transient metabolic mechanism. Given that A–P targets protozoa, we reanalyzed metagenomic microbiome data using a comprehensive reference database that includes protozoan, bacterial, and viral genomes to evaluate the presence of protozoan pathogens in CRC patients.

The complementary metagenomic analysis revealed a strong association between *T. gondii* and CRC. Among 8,425 taxa detected, *T. gondii* emerged as the most discriminatory taxon, with CRC patients exhibiting significantly higher read counts than controls. A dose–response relationship was observed, with cancer risk increasing alongside *T. gondii* read abundance, further supporting the biological plausibility of this association. Elevated *T. gondii* levels were also observed in patients with large adenomas, suggesting a potential role in the progression from premalignant lesions to carcinoma. Notably, prior studies have reported associations between *T. gondii* infection and increased risk of several cancers, including CRC.^20^

These findings support the hypothesis that *T. gondii* may be an underrecognized microbial risk factor for digestive malignancies and that A–P may confer chemopreventive effects by targeting this protozoan pathogen.

### Potential Mechanisms

Atovaquone and Proguanil target protozoan-specific pathways absent in human cells: atovaquone disrupts mitochondrial function by inhibiting the cytochrome bc1 complex of the protozoan electron transport chain^21^, while proguanil—via its active metabolite cycloguanil—inhibits dihydrofolate reductase (DHFR), impairing DNA synthesis in susceptible organisms. The combination has documented activity against a range of protozoan pathogens, including *Toxoplasma gondii*, *Plasmodium* spp., *Pneumocystis jirovecii*, *Babesia microti*, *Cryptosporidium parvum*, *Leishmania* spp., *Entamoeba histolytica*, and *Trichomonas vaginalis*. While often prescribed for malarial prophylaxis, atovaquone, in combination with pyrimethamine is frequently used for the treatment of toxoplasmosis^8^.

Importantly, A–P lacks direct antibacterial or antifungal activity, suggesting that its influence on the gut microbiota is primarily mediated through antiparasitic effects. *T. gondii*, a protozoan widely present in the human environment, is known to chronically colonize the gastrointestinal tract and elicit persistent mucosal inflammation.

In our metagenomic analysis using the SANBAFPH reference database, which includes 4,668 species-level assemblies (4,240 bacterial, 35 protozoan, and 81 fungal species), *T. gondii* emerged as the most differentially expressed in colorectal cancer samples. This finding supports the hypothesis that *T. gondii*, rather than broad microbiome disruption, may underlie the observed chemopreventive association between A–P use and digestive tract cancers.

The observed oncogenic potential of *T. gondii* may be mediated by several mechanisms. Chronic infection in the gastrointestinal mucosa can trigger sustained inflammation, driven by proinflammatory cytokines such as IL-6 and TNF-α. These cytokines activate the NF-κB and STAT3 pathways, promoting epithelial proliferation, immune evasion, and DNA damage^22,23^. Additionally, *T. gondii* infection induces oxidative stress, generating reactive oxygen^24^ and nitrogen species that contribute to mutations in oncogenes (e.g., *KRAS*) and tumor suppressor genes (e.g., *TP53*)^25,26^. *T. gondii* manipulates host cells by inhibiting apoptosis, upregulating antiapoptotic proteins (e.g., Bcl-2) and suppressing p53 activity - allowing mutated cells to survive and accumulate further genetic alterations, further contributing to tumorigenesis.^27^

Interestingly, while our findings suggest an oncogenic role for *T. gondii*, prior studies have reported conflicting evidence of its immunomodulatory effects, including potential antitumor activity^28^. Experimental models suggest that *T. gondii* infection can modulate host immune responses and tumor-associated gene expression in ways that may inhibit certain tumor types^29^. These discrepancies underscore the complexity of host–parasite interactions, which may vary depending on *T. gondii* genotype, infection burden, host immunity, and tumor microenvironment. Future mechanistic studies are warranted to clarify the role of *T. gondii* across different parasite genotypes, tissue types and cancer contexts.

### Broader Implications and A-P as a Preventive Agent

The observed reduction in pancreatic cancer incidence aligns with increasing evidence implicating chronic infections in pancreatic carcinogenesis, as demonstrated with *Helicobacter pylori* and hepatitis viruses^30^. The ability of A-P to reduce cancer risk across multiple digestive organs supports the hypothesis that *T. gondii* may have a broader pathogenic role. Atovaquone, one component of A-P, is a well-established treatment for toxoplasmosis, inhibiting the parasite’s mitochondrial cytochrome b^21^. Its safety profile and established efficacy in treating *T. gondii* infections make it a candidate for further investigation in cancer prevention.

### Diagnostic Potential

Our study also suggests the potential utility of *T. gondii* DNA as a biomarker for CRC detection. Despite inherent noise and nonspecificity in shotgun metagenomic data, *T. gondii* read counts demonstrated promising discriminatory power, with an area under the curve (AUC) of 0.720, comparable to fecal occult blood testing (FOBT). Future diagnostic approaches could employ targeted detection methods, such as PCR using *T. gondii*-specific primers, possibly in association with immunohistochemical assays for parasite antigens or occult blood, to improve specificity and sensitivity. Furthermore, if *T. gondii* contributes to the development of pancreatic as well as colorectal malignancies, fecal detection of the parasite may offer a novel approach for early detection or risk stratification in pancreatic cancer. This also raises the possibility that treatment with antiparasitic agents such as atovaquone–proguanil (A–P) might reduce cancer risk in individuals harboring the parasite.

### Limitations

This study has limitations. The TriNetX data are observational, and while propensity score matching reduced confounding, causality cannot be definitively established. Additionally, the metagenomic dataset is relatively small, and fecal sampling may not fully capture the dynamic interactions within the gut microbiota. The lack of detailed data on A–P dosing and adherence in TriNetX may also influence the accuracy of effect size estimates. While Centrifuge allowed detection of protozoa using a comprehensive reference database, its read-level outputs are not normalized for genome size or sequencing depth, limiting precise abundance estimation. However, consistent DNA extraction and sequencing protocols across samples, and the averaging of species-level read counts across all samples from each individual, help support valid semi-quantitative comparisons. Lastly, although *T. gondii* showed a robust association with CRC, we did not assess associations with other digestive cancers beyond pancreatic malignancies. Moreover, the mechanism by which *T. gondii* may promote tumorigenesis—and how A–P may counteract these effects—remains to be experimentally validated in cellular or animal models.

### Future Directions

Prospective studies and randomized controlled trials are needed to validate our findings, establish causality, and explore the potential of A-P for cancer prevention. Further mechanistic research is essential to elucidate the role of *T. gondii* in digestive carcinogenesis, particularly its interactions with the host immune system. In addition, expanded metagenomic analyses across diverse populations could enhance understanding of regional differences in infection prevalence and cancer risk. Investigations into optimized diagnostic tools targeting *T. gondii* DNA or antigens could also advance non-invasive CRC screening strategies.

## Conclusion

In this retrospective cohort analysis, A-P use was associated with a reduced risk of digestive cancers, notably colorectal and pancreatic, and *T. gondii* DNA was strongly associated with CRC. These findings suggest that *T. gondii* may be an overlooked microbial risk factor for digestive cancers and that targeting *T. gondii* with antiparasitic therapy may represent a novel avenue for cancer prevention. Prospective studies and mechanistic research are warranted to explore the therapeutic and diagnostic implications of targeting *T. gondii* in digestive oncology.

## Data Availability

TriNetX epidemiologic data are accessible via the TriNetX consortium. Metagenomic sequencing data are accessible in the European Nucleotide Archive under accession number PRJEB6070."

## Data Sharing Statement

The metagenomic dataset (PRJEB6070) is publicly available through the European Nucleotide Archive. The TriNetX data used in this study are not publicly available.

## Authors Contribution

Conceptualization: AI, EMe, Ema

Methodology: AI, EMe

Investigation: AI, SI, EMe

Writing - original draft: AI

Writing - review & editing: AI, AW, SI, SA, SV, EMa, EMe

AI and EMe had full access to all the data and takes responsibility for the integrity of the data and accuracy of the analysis. All authors contributed to drafting and revising the manuscript

## Preprint

A preprint version of this study has been made available on medRxiv^31^.

## Generative AI statement

ChatGPT (version 4o, OpenAI) was used to assist with language refinement for improved clarity and flow, and to provide support with coding tasks related to data analysis scripts. All content was reviewed and verified by the authors to ensure accuracy and integrity.

## Abbreviations

A-P: Atovaquone-Proguanil
BMI: Body mass Index
CI: Confidence Interval
CRC: colorectal cancer
DM: Diabetes Mellitus
FOBT: Fecal occult blood test
HR: Hazard ratio
SMD: Standardized Mean Difference

